# Formal Statistical Replication Analysis in Lung Cancer Genome-Wide Association Studies

**DOI:** 10.1101/2025.10.02.25337130

**Authors:** Yung-Han Chang, Jinyoung Byun, Bryan R. Gorman, Rayjean J. Hung, James D. McKay, Christopher I. Amos, Saiju Pyarajan, Arjun Bhattacharya, Ryan Sun

**Affiliations:** Department of Biostatistics, University of Texas MD Anderson Cancer Center UTHealth Houston Graduate School of Biomedical Sciences, Houston, TX, USA; Institute for Clinical and Translational Research, Baylor College of Medicine, Houston, TX, USA; Department of Medicine, Section of Epidemiology and Population Sciences, Baylor College of Medicine, Houston, TX, USA; University of New Mexico Comprehensive Cancer Center, Albuquerque, NM, USA; Center for Data and Computational Sciences (C-DACS), VA Cooperative Studies Program, VA Boston Healthcare System, Boston, MA, USA; Dalla Lana School of Public Health, University of Toronto, Toronto, Ontario, Canada; Prosserman Centre for Population Health Research, Lunenfeld-Tanenbaum Research Institute, Sinai Health System, Toronto, Ontario, Canada; Section of Genetics, International Agency for Research on Cancer, World Health Organization, Lyon, France; Dan L Duncan Comprehensive Cancer Center, Baylor College of Medicine, Houston, TX, USA; Department of Medicine, Brigham and Women’s Hospital, Harvard Medical School, Boston, MA, USA; Department of Epidemiology, University of Texas MD Anderson Cancer Center, Houston, TX, USA; Institute for Data Science in Oncology, University of Texas MD Anderson Cancer Center, Houston, TX, USA

## Abstract

Dozens of genome-wide association studies (GWAS) have identified thousands of single nucleotide polymorphisms (SNPs) associated with lung cancer risk. However, it remains challenging to translate these findings to clinical insights. One well-known obstacle is the large amount of type I error attached to GWAS; attempted solutions such as setting a *p*-value threshold across multiple cohorts or looking for small meta-analysis *p*-values have only somewhat reduced false positive findings. In contrast, here we advocate for a statistical model-based replication analysis. We first demonstrate that a formal statistical test for the replication composite null hypothesis - i.e. that the regression coefficient of a SNP falls in the same direction in multiple cohorts simultaneously - can curate a smaller, higher-quality list of significant SNPs than common alternatives. In two-way simulations, the false discovery rate (FDR) of model-based replication analysis is 6.4 times lower than that of meta-analysis with a *p <* 10^−8^ threshold. In three-way replication analysis, 9.8% of the International Lung Cancer Consortium GWAS significant SNPs are replicated for squamous cell lung cancer while 33.8% are replicated for lung adenocarcinoma. Finally, we construct polygenic risk scores (PRSs) and find the replication-based PRS achieves virtually identical performance to a GWAS-significant PRS while using 87.3% fewer variants. Thus, formal model-based replication analysis can greatly reduce spurious findings while still identifying important variants, allowing for more robust and more efficient translation of GWAS results.

## Introduction

Lung cancer is the leading cause of cancer-related mortality in both men and women [1, 2, 3, 4]. While tobacco use is the primary risk factor, individuals who have never smoked or had significant environmental exposures may still face an increased risk due to inherited genetic factors [1, 2, 5, 6]. The heritability of lung cancer has been estimated at 15-18% [7].

Dozens of lung cancer genome-wide association studies (GWAS) have been performed in varied settings and diverse populations, and this approach still remains highly popular, with many substantial recent and ongoing efforts [1, 8, 9]. However, despite researchers identifying thousands of genetic variants associated with lung cancer risk over the past two decades [8, 10, 11, 12], there still exists a significant portion of missing heritability, and clinical applications of GWAS findings remain limited. Such a trend suggests that the continued unearthing of new associations may not be producing actionable insights.

One substantial, well-understood obstacle of the GWAS approach is the large amount of type I error it produces. Because many non-functional single nucleotide polymorphisms (SNPs) lie in linkage disequilibrium (LD) with true causal variants, GWAS highlights many unimportant variants. While some highly significant variants are consistently identified across studies, a much larger number are often falsely emphasized as well, as we will show. Such unimportant noise greatly hinders downstream translational efforts such as building polygenic risk scores (PRSs) or developing targeted therapies [1, 8, 9, 13, 14, 15].

Efforts to prune inconsequential SNPs, such as requiring a certain *p*-value threshold in multiple datasets or using significant meta-analysis results that integrate different cohorts, have only somewhat improved the quality of findings [16]. Although the value of replication is well-recognized, the lack of replicability has been consistently cited as a major challenge facing genetics research. The number of lung cancer replication studies remains very small in proportion to the number of new GWAS or GWAS meta-analysis investigations [2]. One possible reason for the scarcity of replication literature is the lack of robust, systematic, and interpretable methods for testing the replication null hypothesis [17, 18].

Here, we advocate for and perform formal statistical model-based replication analysis in lung cancer GWAS cohorts. Our overall goal is to provide an interpretable and reliable framework for reducing false positives while still highlighting important variants. This framework can be straightforwardly extended to other phenotypes.

We first demonstrate how to utilize a version of the popular empirical Bayes two-group model to test the replication composite null hypothesis [19]. This null hypothesis is rarely explicitly stated in GWAS studies. However, the definition of the replication composite null - that a SNP regression coefficient is non-zero and falls in the same direction across multiple cohorts simultaneously - truly matches the scientific purpose of a replication study (see Figure 1) [18].

**Figure 1:**
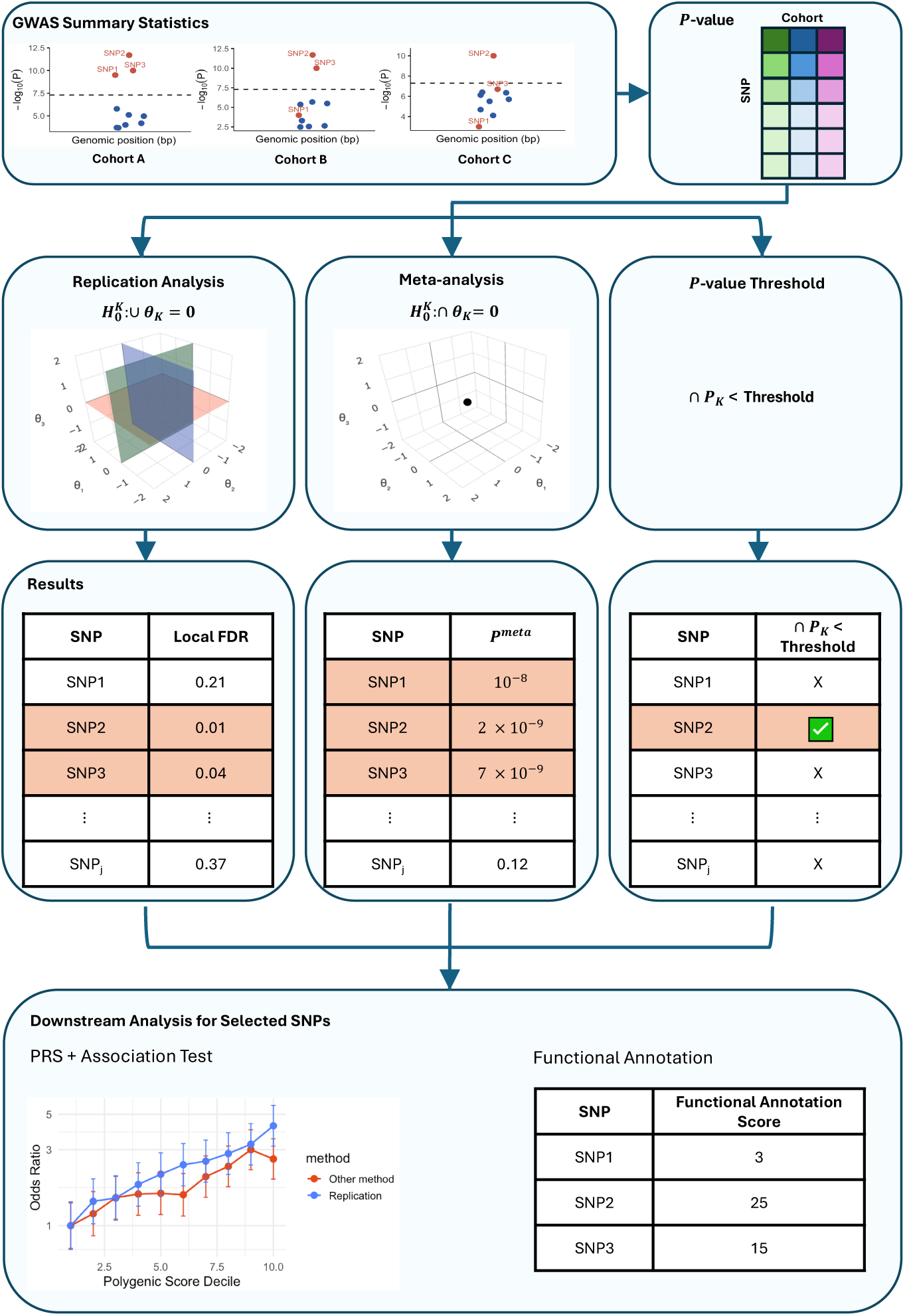
*Conceptual comparison of replication analysis, meta-analysis, and p-value threshold approach.* Replication analysis tests the composite null, requiring SNPs to show consistent associations across all cohorts (e.g., SNPs 2 and 3). Meta-analysis instead tests the global null, so it can reject based on signal in a single cohort (e.g., SNP 1). The *p*-value threshold method requires a SNP to pass a fixed cutoff in every cohort, which can miss true signals (e.g., SNP 3 in Cohort C with a p-value barely greater than 5 × 10^−8^). The three-dimensional schematics illustrate that under meta-analysis the global null is the single point at the origin (0,0,0), whereas a composite null (shaded planes) includes all points where at least one parameter is zero; in three dimensions this composite null region is much larger than the global null point. This geometric interpretation explains why the empirical Bayes replication test offers stronger protection against false discoveries as the number of cohorts (dimensions) increases. We further construct PRSs and conduct functional annotation analysis to test the quality of selected SNPs.

In realistic simulations, model-based replication shows a 6.4-fold lower false discovery rate (FDR) compared to meta-analysis (using a significance level of *p <* 10^−8^) and improved power by 19.1%. The proposed method also has the added advantage of adaptivity to different strengths of signals across different datasets. Additionally, model-based replication exhibits increased relative power and FDR calibration as the number of cohorts increases. Across three large lung cancer GWAS cohorts [8, 20, 21], we find that a PRS based on replication-based SNPs shows virtually identical performance to a GWAS-significant PRS while using 87.3% fewer SNPs.

Our findings indicate that model-based replication analysis can robustly filter out false positives and curate a more concise list of important SNPs from multiple GWAS. This filtering of false positives, and retention of true positives, provides higher-quality findings for follow-up translational research and improves the reproducibility of GWAS investigations. In certain cases, model-based replication testing can reduce the number of putatively significant findings by an order of magnitude. Additionally, while this work focuses on lung cancer, the approaches are broadly generalizable to other phenotypes.

## Results

### Overview of model-based replication analysis

To assess replicability across cohorts, we employed model-based replication analysis, which tests the composite null hypothesis [19, 22]. In this framework, a SNP is considered null if it shows no effect in at least one cohort or if effect directions differ across cohorts, making rejection more stringent than the meta-analysis approach for integrating multiple GWAS [1, 9, 13, 14].

Let *θ_jk_* denote the cohort-specific regression coefficient for SNP *j* in cohort *k*, for *k* = 1*, …, K*. We define the replication composite null as

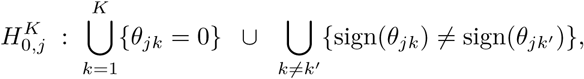

Replication testing was implemented using a multidimensional empirical Bayes two-group approach [18], with density estimation based on the conditionally symmetric Gaussian mixture model (csmGmm) [19]. This density model has been shown to provide robust operating characteristics and, crucially, offers interpretability guarantees by preventing frequentist-Bayesian contradictions. Within this framework, replication testing calculates the empirical Bayes local false discovery rate (lfdr) [23] for each SNP and uses it as a decision rule to identify variants that replicate across cohorts. See Figure 1 and Methods for further details.

To demonstrate the benefits of model-based replication analysis, we first conducted simulations varying mean effect sizes and proportions of causal SNPs in two- and three-cohort settings, and we then applied the method to three large cancer GWAS summary datasets, comparing its performance against the *p*-value threshold approach (selecting SNPs by a fixed threshold across all cohorts) and meta-analysis. We further assessed the quality of findings by constructing PRSs and conducting functional annotation analyses.

### Realistic simulations show that replication analysis properly calibrates FDR while maintaining power

The simulation studies show that model-based replication analysis offers (i) robust control of false discoveries, (ii) improved statistical power, and (iii) adaptivity to different signal strengths across datasets. Additionally, (iv) its advantages become greater as the number of datasets increases.

The first set of simulations shown in Figure 2A, 2C, and 2E demonstrate (i) that model-based replication analysis can protect nominal FDRs across a range of sparsity and effect size settings. When the proportion of causal variants is 0.02%, there are roughly 1,400 causal SNPs (Figure 2A). When the proportion is 1%, there are 700,000 causal SNPs (Figure 2E). In each case, the model-based approach robustly constrains the number of false discoveries. This robust performance holds when the signals are very weak (causal summary statistic means of 2) and very strong (causal summary statistic means of 5+).

**Figure 2:**
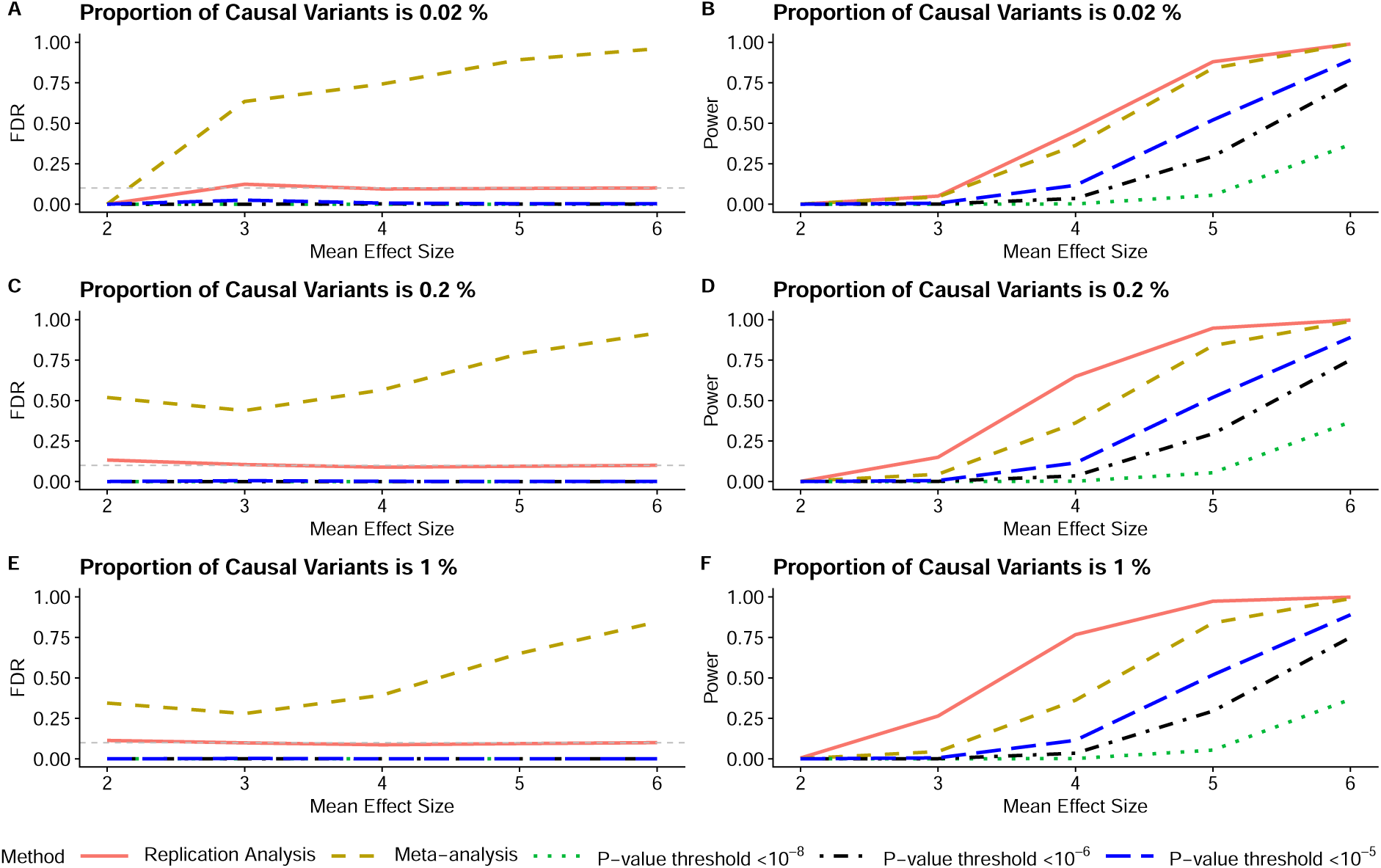
*Two-way replication analysis simulation evaluating the impact of varying mean effect sizes and proportions of causal variants.* Each line corresponds to a different analysis method. For the meta-analysis, significance was defined at *p <* 1 × 10^−8^. Each *p*-value threshold line corresponds to the approach of selecting only SNPs passing the given significance cutoff (*p <* threshold) in both datasets. The x-axis shows the mean effect size, while the y-axis displays either FDR (left panels) or statistical power (right panels). The gray horizontal line in the left panels marks the nominal FDR = 0.1.

In contrast, meta-analysis demonstrates extremely large numbers of false discoveries, as would be expected, given that it does not test the replication null hypothesis. The meta-analysis significance level of *p <* 1 × 10^−8^ is chosen in part to keep its empirical FDR reasonable. A less stringent significance level brings the FDR to near 1. On the other hand, the *p*-value threshold approach of looking for SNPs under a threshold in both datasets is extremely conservative, with observed FDR near 0 in all settings. This poor performance holds regardless of the threshold chosen (*p <* 10^−8^ down to *p <* 10^−5^).

Figures 2B, 2D, and 2F further show (ii) that model-based replication analysis offers good power to detect truly replicated SNPs. The model-based replication approach detects substantially more signals than the threshold approaches, which is expected given the extreme conservativeness of the threshold methods. The empirical Bayes model also shows slightly more power than meta-analysis, even though meta-analysis makes many more false discoveries (see Supplementary Appendix A for more FDR and power simulations across different settings).

Additionally, the suggested model shows larger power advantages when mean effect sizes are small to moderate, e.g. below 5 on the x-axis. This behavior is partly due to (iii) the adaptiveness of the test. Methods such as the threshold approach or meta-analysis rely on a fixed significance level such as 1 × 10^−8^, and if the signals are not sufficiently strong, power will be very low. However, the empirical Bayes model can adapt to low overall signal strength and still identify SNPs with relatively strong associations across multiple datasets.

Figure 3 displays key differences between two-way and three-way replication analysis, especially (iv), with many advantages of model-based replication testing increasing in the three dimensional setting. In particular, the empirical Bayes model shows much better protection of false discoveries than meta-analysis, as expected since the composite null expands relative to the global null in three dimensions compared with two (Figure 1). The model-based replication test also continues to show much better adherence to the nominal FDR than the threshold approaches, which make almost no discoveries when signals are small to moderate.

**Figure 3:**
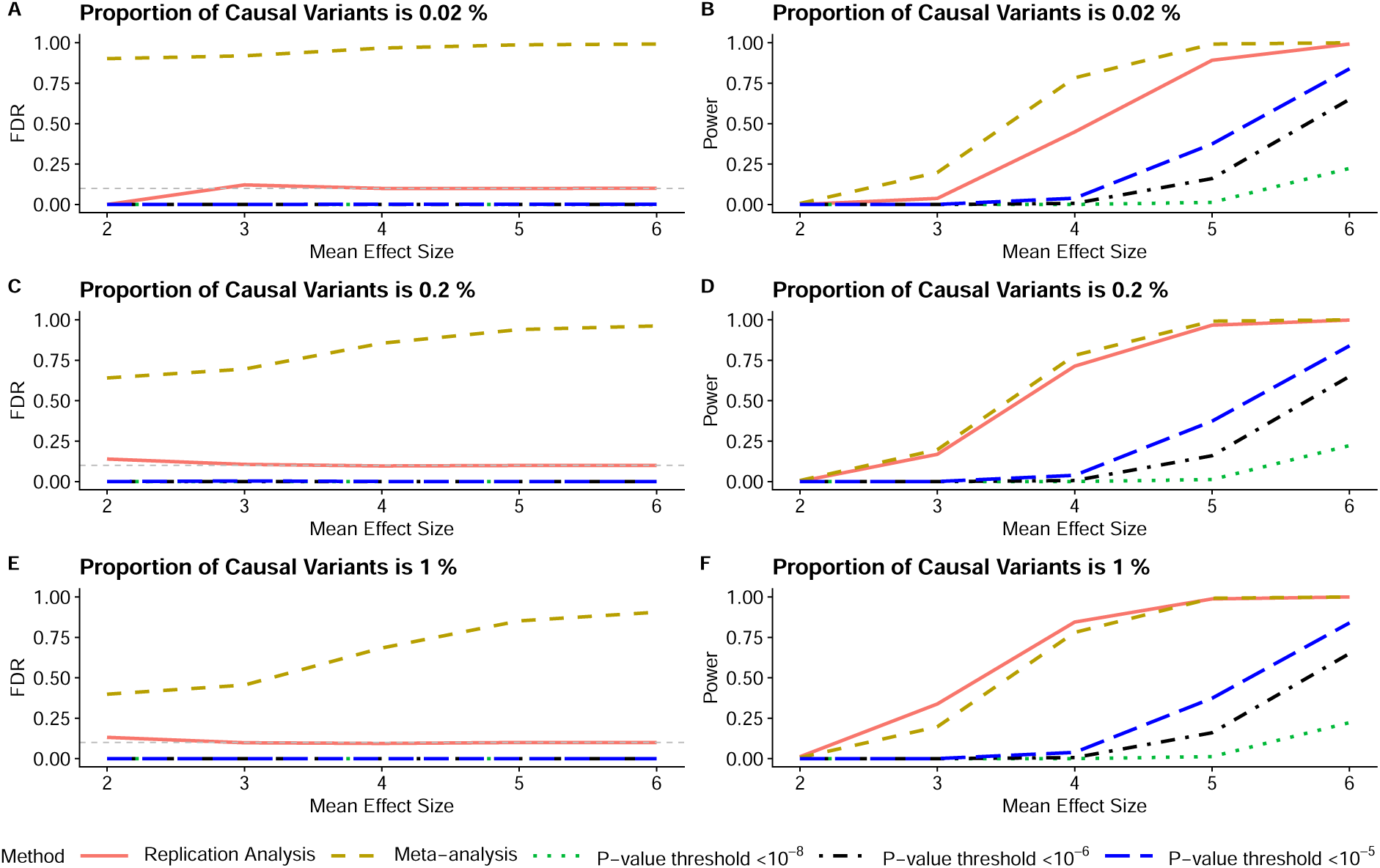
*Three-way replication analysis simulation evaluating the impact of varying mean effect sizes and proportions of causal variants.* The x-axis is the mean effect size, while the y-axis is either statistical power or the FDR, depending on the panel. The advantages of model-based replication analysis are more extreme in three dimensions than in two dimensions. The gray horizontal line in the left panels marks the nominal FDR = 0.1.

The model-based replication approach also achieves greater power than the threshold approach in three dimensions. This is expected, since reaching a significance threshold across three datasets is more difficult than across two, even for true signals. Meta-analysis can show slightly more power in three-way settings, but it is clearly an invalid test given the extremely large numbers of false discoveries.

Figure 3 succinctly illustrates the dangers of using meta-analysis or the threshold approach to identify high-quality SNPs with large numbers of datasets. As additional cohorts are included, meta-analysis identifies an increasing number of false positives, because the global null becomes more different from the composite null as the number of dimensions increases. Additionally, as the number of datasets grows, the threshold approaches impose increasingly stringent criteria, yielding only the most obvious findings. Further simulation settings are available in the Supplementary Materials.

### Two-way replication analysis curates higher-quality SNPs than meta-analysis in real data

While our primary focus is on the three-way replication analysis of the International Lung Cancer Consortium (ILCCO), Million Veteran Program (MVP), and UK Biobank (UKB) datasets, we first briefly present two-way replication results, which follow patterns similar to the simulation studies above. Specifically, Extended Data Figure 1 presents Manhattan plots of the replication analysis results on the two largest datasets (ILCCO and MVP) for overall lung cancer, LUAD, and LUSC. Additional two-way results with the UK Biobank can be found in Supplementary Appendix B.

In total, there are 3,736 SNPs with standard GWAS *p <* 5 × 10^−8^ for overall lung cancer in the ILCCO dataset. Of the 3,736 SNPs, 12.8% are identified as significant in the model-based replication analysis, and 9.9% are identified as significant under the *p*-value threshold approach at 1 × 10^−8^. However, 81.5% are declared significant in meta-analysis; many of these SNPs correspond to the green dots at the bottom of Extended Data Figure 1A. Thus, meta-analysis is ineffective for curating higher-quality SNPs in overall lung cancer using these two datasets, as it largely reproduces the set of originally significant ILCCO SNPs.

For LUAD, there are 1,160 SNPs with standard GWAS *p <* 5 × 10^−8^ in the ILCCO data (Extended Data Figure 1B). Of the 1,160, 40.4% are significant in the model-based replication analysis, and 4.1% are significant under the *p*-value threshold approach at 1 × 10^−8^. By contrast, 89.7% are significant by meta-analysis.

We can see that the overall lung cancer and LUAD examples reinforce themes (i) and (ii) from the simulations. The model-based replication analysis appears to offer much better protection of false positives than meta-analysis while also offering much more power than the threshold approach. The model-based analysis can also adapt to the lower levels of signal in LUAD (where the sample size is smaller than for overall lung cancer) and identify a much larger proportion of replication SNPs.

The LUSC example of Extended Data Figure 1C is noteworthy because it shows how model-based replication testing can also identify high-quality variants that may be overlooked amid thousands of significant SNPs. For example, a locus in the 9p21 region of chromosome 9 is uniquely identified by the model-based analysis. Given the simulations (Figure 2) showing that model-based replication analysis can have more power than meta-analysis, it is reasonable to believe that this locus is an important finding. The ability to identify SNPs with moderate signals across multiple datasets is an important complement to filtering out spurious associations.

### Three-way replication analysis reveals additional loci missed by meta-analysis

We next present the results of three-way replication analysis in the ILCCO, MVP, and UKB datasets. There are substantial differences in signal size between the three datasets, with smallest *p*-values for overall lung cancer being *p* ≈ 10^−100^ in the ILCCO dataset, *p* ≈ 10^−25^ in MVP, and *p* ≈ 10^−7^ in the UKB. Replication results are shown overlaid on the original GWAS in Figure 4 and separately in Extended Data Figure 2, with selected summaries provided in Table 1 (additional results available in Supplementary Appendix C).

**Figure 4:**
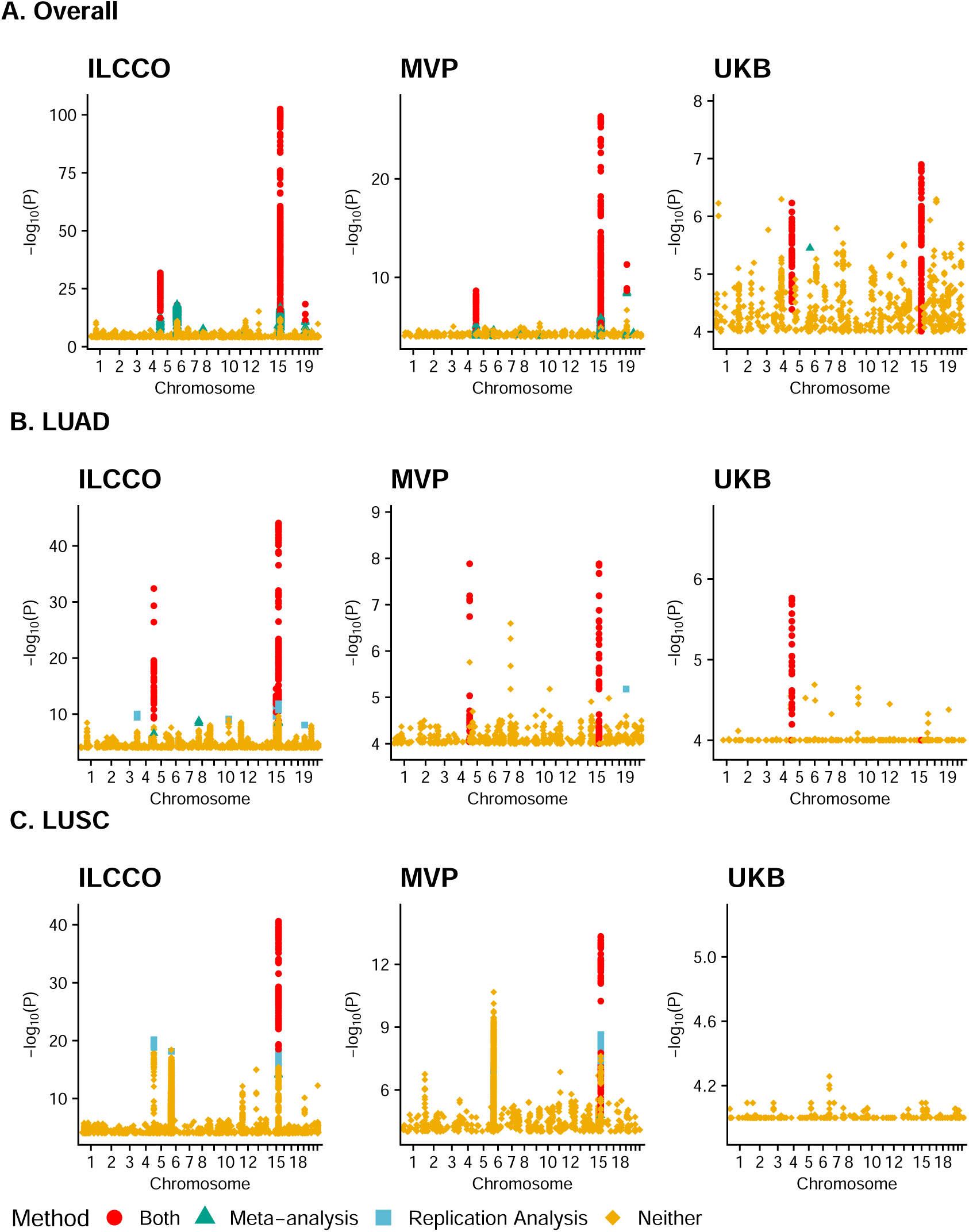
*Manhattan Plots of Lung Cancer GWAS Summary Statistics by Subtype and Cohort.* The y-axis is the −*log*_10_(*P*) value from the original GWAS of each subtype in each cohort. SNPs are colored red if significant in both model-based replication analysis and meta-analysis, green if significant only in meta-analysis, and blue if significant only in model-based replication analysis. Only SNPs with original GWAS *p <* 10^−4^ are displayed, and the exact counts of significant SNPs are provided in Table 1.

**Table 1:** *Summary of significant SNPs identified by GWAS, model-based replication analysis, meta-analysis, and p-value threshold approaches in ILCCO, MVP, and UKB.* For ILCCO and MVP, percentages represent the proportion of GWAS-significant SNPs identified by each method. No genome-wide significant SNPs were detected in UKB. Raw counts are provided in the Total column.

|  |  | ILCCO | MVP | UKB | Total |
| --- | --- | --- | --- | --- | --- |
| Overall | Total # Significant ($p < 5 \times 10^{-8}$) | 3736 | 392 | 0 | 4128 |
|  | Total # Significant in Replication | 12.3% | 98.9% |  | 461 |
|  | Total # Significant in Meta-Analysis | 37.9% | 99.5% |  | 1472 |
| | Total # Significant using Threshold $10^{-6}$ | 0.7% | 6.6% | | 26 |
| | Total # Significant using Threshold $10^{-5}$ | 3.2% | 30.9% | | 121 |
|  | Total # Significant in Rep + Meta | 12.3% | 98.9% |  | 461 |
| LUAD | Total # Significant ($p < 5 \times 10^{-8}$) | 1160 | 56 | 0 | 1216 |
|  | Total # Significant in Replication | 33.8% | 96.4% |  | 392 |
|  | Total # Significant in Meta-Analysis | 31.9% | 96.4% |  | 371 |
| | Total # Significant using Threshold $10^{-6}$ | 0 | 0 | | 0 |
| | Total # Significant using Threshold $10^{-5}$ | 2.5% | 51.8% | | 29 |
|  | Total # Significant in Rep + Meta | 31.1% | 96.4% |  | 361 |
| LUSC | Total # Significant ($p < 5 \times 10^{-8}$) | 2555 | 473 | 0 | 3028 |
|  | Total # Significant in Replication | 9.8% | 20.7% |  | 250 |
|  | Total # Significant in Meta-Analysis | 7.5% | 11.2% |  | 189 |
| | Total # Significant using Threshold $10^{-6}$ | 0 | 0 | | 0 |
| | Total # Significant using Threshold $10^{-5}$ | 0 | 0 | | 0 |
|  | Total # Significant in Rep + Meta | 7.4% | 11.2% |  | 189 |

Figure 4A illustrates (iii), the data-adaptive behavior of the model-based replication analysis. Although there are no traditional genome-wide significant signals in the UKB dataset for overall lung cancer, the empirical Bayes model adapts to the low overall signal level and still identifies 461 SNPs with relatively small *p*-values that are replicated in the ILCCO and MVP datasets. These SNPs fall in the well-chronicled chromosome 5p15.33 and chromosome 15q25 regions, corresponding to the *TERT* [24, 25] and nicotine receptor loci [26, 27, 28], respectively, as expected.

Figures 4B, 4C, and Extended Data Figure 2 further demonstrate that model-based replication testing can identify important SNPs that meta-analysis misses. For example, the chromosome 3q28 locus is identified in LUAD only by model-based replication testing, not by meta-analysis (Figures 4B, Extended Data Figure 2C, 2D). Additional loci uniquely identified by model-based replication testing include chromosome 5p15.33 in LUSC (Figures 4C, Extended Data Figure 2E, 2F) and chromosome 6p21.33 for overall lung cancer and LUSC (Extended Data Figure 2A, 2B, 2E, 2F). The chromosome 6 region in particular has received heightened scrutiny lately due to immune-related processes implicated in lung cancer [29, 30].

Table 1 provides additional evidence of (iv), illustrating how three-dimensional settings can exacerbate differences between model-based replication analysis, meta-analysis, and the threshold approaches. We see that for overall lung cancer, only 12.3% of the original ILCCO GWAS-significant SNPs are identified as significant by model-based replication testing, while 37.9% are identified as significant by meta-analysis. Thus, meta-analysis identifies approximately three times as many SNPs, and previous simulations suggest that many of these are likely false positives. For both LUAD and LUSC, model-based replication analysis and meta-analysis make similar numbers of discoveries. However, for both subtypes, the threshold approach at *p <* 1×10^−6^ finds no significant SNPs, whereas the replication analysis finds hundreds of significant SNPs. This dramatic difference in the number of findings further emphasizes the lack of power for the threshold approach.

The most significant SNPs identified in the three-way replication analysis for each lung cancer subtype are presented in Extended Data Table 1. We can see that generally the ILCCO data provides the strongest evidence of association, and the UKB data provides the least. The full list of significant findings, along with all results, pipelines, and intermediate files used in this analysis, are available at locations described in the Data Availability.

### Replication-based PRS achieves virtually identical performance with far fewer variants

We constructed two PRSs for overall lung cancer: (a) a GWAS-significant PRS based on ILCCO variants with *p <* 5 × 10^−8^ (3,736 SNPs), and (b) a two-way replication PRS constructed from variants significant in both ILCCO and MVP (478 SNPs) by PRSice [31] (see Methods). Replication-based PRS achieved virtually identical performance to GWAS-significant PRS while requiring 87.3% fewer genetic variants for overall lung cancer. The AUC was 0.698 for the replication-based PRS and 0.699 for the GWAS-significant PRS; both significantly outperformed the covariate-only model (DeLong Test: replication *p* = 5.65 × 10^−7^; GWAS *p* = 3.59 × 10^−7^). The adjusted odds ratio (OR) for the top 10% versus bottom 10% PRS was 1.97 for the replication-based PRS and 1.88 for the GWAS-significant PRS (Figure 5). Case rates showed similar separation, with 1.00% vs. 0.51% for the replication PRS (top 10% vs. bottom 10%) and 0.98% vs. 0.53% for the GWAS PRS.

**Figure 5:**
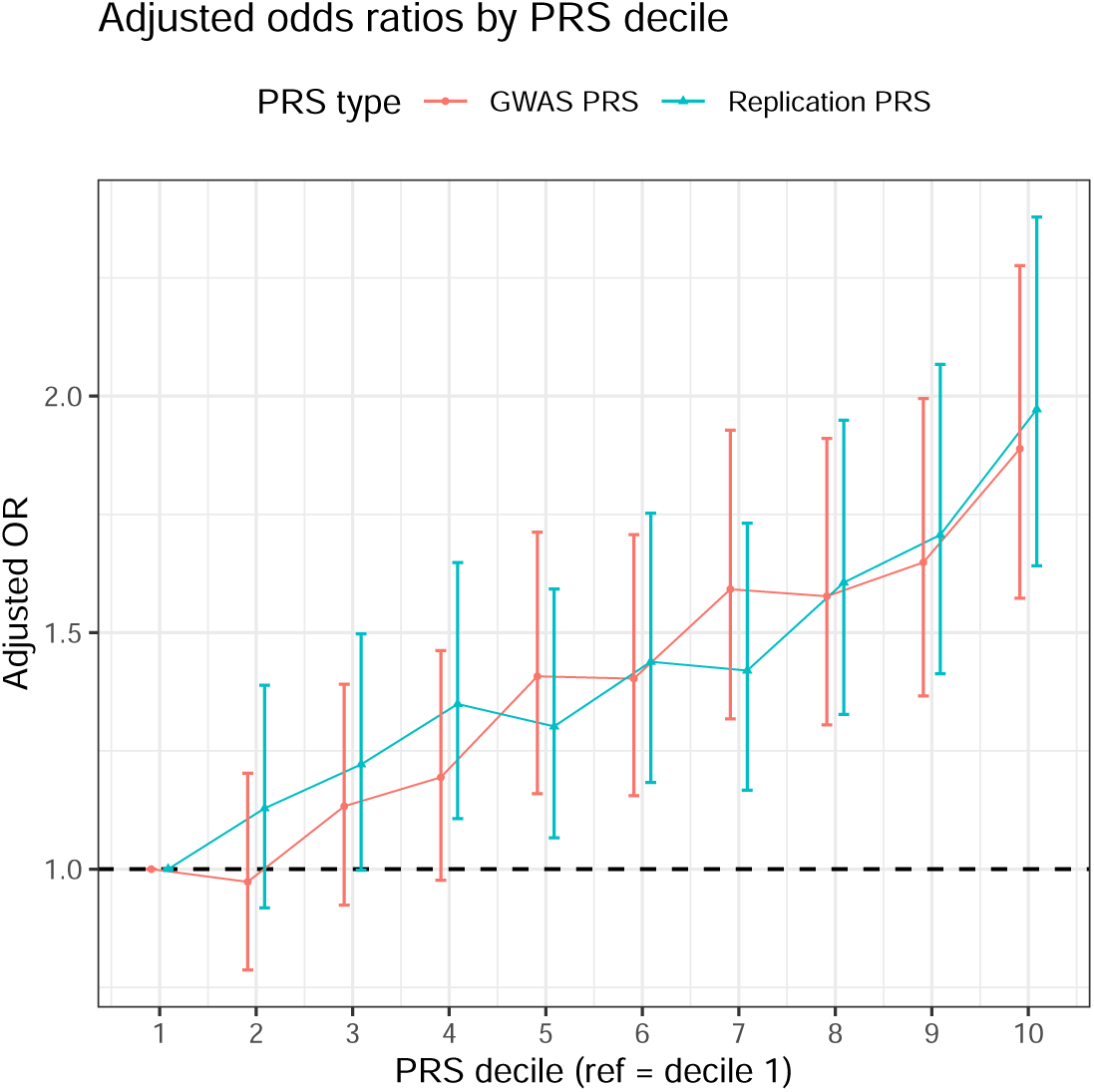
*PRS stratification of overall lung cancer risk in UK Biobank.* Adjusted ORs with 95% CIs are shown across PRS deciles (reference = decile 1). Scores were built using PRSice.

### Replication analysis and meta-analysis identify comparable functionally relevant SNPs

We further investigated whether the quality of findings from replication analysis is comparable to the quality from meta-analysis in a real-data application. While it is generally not possible to label the true causal SNPs in real data, we used genome-wide functional annotation data from Functional Annotation of Variants Online Resources (FAVOR) as a proxy. Specifically, we aligned FAVOR data with replication results at a well-studied locus on chromosome 15q25.1 harboring nicotinic acetylcholine receptor genes [26, 27, 28]. We focused on LUSC, given that the numbers of findings at this locus were reasonably similar between model-based replication analysis (234) and meta-analysis (191).

Figure 6 shows how two FAVOR functional annotation scores range for SNPs identified as significant by model-based replication analysis and meta-analysis. Model-based replication analysis identified 13 (5.6%) SNPs with medium or higher conservation scores and 34 (14.5%) with medium or higher epigenetic activity scores, whereas meta-analysis identified 9 (4.7%) and 32 (16.8%) such SNPs, respectively. Thus, the proportion of SNPs with high functional relevance is similar between the two approaches.

**Figure 6:**
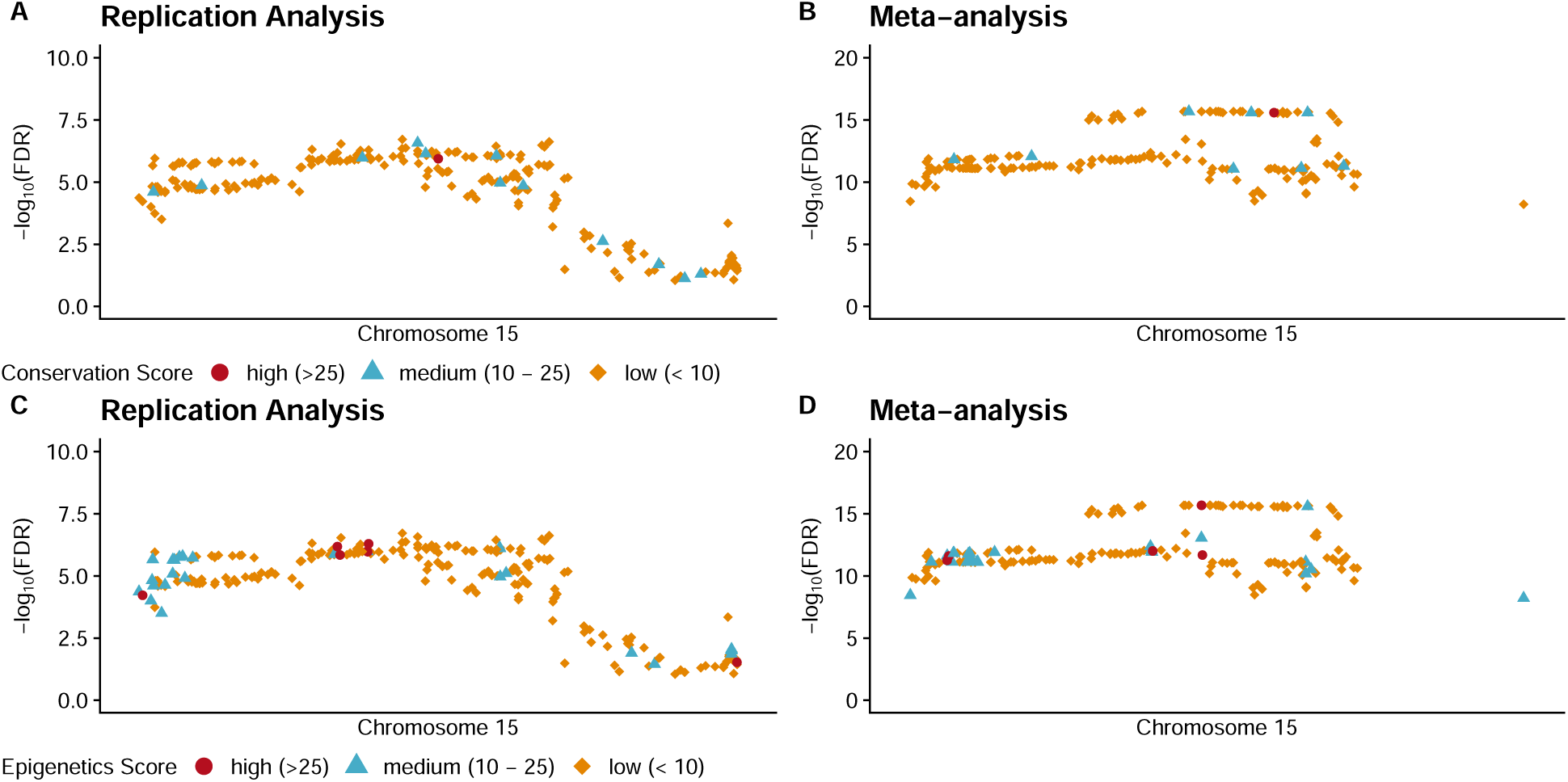
*Functional annotation of variants identified in the three-way real data analysis for LUSC.* The x-axis is genomic position and the y-axis is −*log*_10_(*FDR*). The first column displays results from the replication analysis, and the second column shows results from the meta-analysis. The top row presents conservation scores, while the bottom row presents epigenetic scores. We categorize the scores as follows: scores larger than 25 are considered high, scores between 10 and 25 are medium, and scores less than 10 are low.

Annotation scores represent only one approach to assessing functionality and are not specific to lung cancer. However, these results suggest that, as previously seen, model-based replication analysis offers power comparable to meta-analysis for detecting important variants. Its main advantage remains the ability to substantially reduce the number of putatively significant GWAS findings. By filtering out false positives early, model-based replication analysis ensures that subsequent annotation and translational research efforts are focused on biologically relevant variants.

## Discussion

This work advocates for and conducts an interpretable and statistically principled model-based replication analysis of lung cancer GWAS results. Specifically, we have demonstrated how an empirical Bayes two-group model can be applied to formally test the replication composite null hypothesis in a straightforward manner. This approach improves control of false discoveries and offers greater statistical power compared to commonly used approaches such as the *p*-value threshold approach or meta-analysis. Furthermore, the model-based replication method adapts effectively to the strength of signal in the GWAS summary statistics and can show improved performance as the number of datasets increases.

Simulation studies show that model-based replication analysis consistently maintains effective FDR control while preserving power across different simulation settings, whereas meta-analysis yields inflated false discoveries and the *p*-value threshold approach is overly conservative with little power. Applied to three large lung cancer cohorts, our method demonstrates that many genome-wide significant SNPs could not be replicated, underscoring the prevalence of false positives in many traditional GWASs, while also identifying functionally relevant loci such as the chromosome 6 region missed by meta-analysis. This locus has been linked to inflammatory processes driving lung cancer in previous clinical studies [29, 30], reinforcing the biological plausibility of the replication-based findings. Replication-based PRS further demonstrates the efficiency of the approach, achieving nearly identical predictive performance to a GWAS-significant PRS while requiring almost 90% fewer variants.

With the growing availability of large-scale biobanks and other GWAS, it is increasingly important to integrate their results in a robust manner [32, 33, 34]. For example, thousands of SNPs have been associated with different lung cancer histologies across diverse populations, yet it remains unclear which should be prioritized. The large amount of false positives can substantially hinder clinical efforts such as PRS construction or development of new therapies [35, 36, 37, 38, 39, 40].

We note that some other statistical models have been proposed for assessing replicability, although few have seen widespread adoption. For example, the repfdr, MAMBA, and SCREEN methods are similarly Bayesian frameworks, however they do not offer interpretability guarantees of our suggested approach [41, 42, 43], which can lead to discrepancies between frequentist summary statistics and Bayesian replication findings. They may also require additional preprocessing steps such as SNP pruning.

We also acknowledge several limitations to our work. One limitation is that, due to the sample size of the available GWAS summary statistics, we focus only on individuals of European ancestry. Previous studies have shown that single-ancestry GWAS can be limited by population-specific LD structures, [44, 45, 46]. However, this challenge also presents an opportunity, as an important future direction is to extend model-based replication across diverse ancestries. Utilizing the suggested framework across different ancestries may further remove noise findings that are attributable to LD, since the noise SNPs linked by LD will be different in different ancestries. Another limitation is that we did not account for differences in reference panels used across the GWAS summary statistics. Variability in reference panels can impact imputation quality, LD estimation, and, *p*-values and downstream analyses [47, 48]. Finally, another direction for future work is exploring the viability of testing for replication in only a subset of datasets. For example, if there are four datasets of interests, one might focus on identifying SNPs that replicate in at least three, which could balance stringency with greater power.

## Materials and Methods

### Lung cancer datasets

The ILCCO GWAS is the largest lung cancer GWAS, with 29,266 cases and 56,450 controls of European ancestry. Subtype specific analyses for LUAD and LUSC include 11,273 and 7,426 cases, respectively. Details about the study and its summary statistics are available in the literature [8].

The MVP GWAS is a recent study of 10,398 lung cancer cases and 62,708 controls of European ancestry that do not overlap with previous lung cancer cohorts. Subtype specific analyses for LUAD and LUSC include 2,019 and 1,475 cases, respectively. Details about the study and its summary statistics are also available in the literature [20].

We further performed a GWAS in the UK Biobank on 2,404 cases and 330,018 controls of white, British ancestry [21]. The LUSC subtype analysis included 489 cases, and the LUAD subtype analysis included 862 cases. We used the REGENIE [49] default pipeline to generate summary statistics, setting a minor allele frequency threshold of 0.00005. We corrected for the same covariates as the ILCCO analysis, using age, gender, and the first 10 ancestry principal components (PCs) in the regression model.

Data harmonization was applied to keep only the variants shared in common by all three datasets. We also flipped effect directions if necessary to ensure that effect alleles and reference alleles were constant across studies. After all data cleaning steps, there were 7,335,666 common SNPs in the analysis for overall lung cancer, 7,325,055 for LUSC, and 7,327,929 for LUAD.

### Overview of replication analysis through testing composite null hypotheses

In general, GWAS summary statistics from two cohorts A and B are generated from logistic regression models of the form

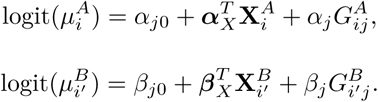

Here, for each subject *i* = 1*, …, n_A_* in cohort A, the 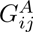 is the genotype at SNP *j*, the lung cancer outcome is 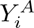, and there is a vector of *p* additional covariates 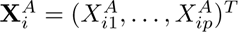 such as principal components of the genotype matrix. Then 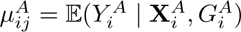. All the same definitions carry over to the *i*^′^ = 1*, …, n_B_* subjects in cohort B. As an example, cohort A could be ILCCO and cohort B could be MVP. For a third cohort C, e.g. the UK Biobank, we would fit a third model of the same form where *γ_j_*is the regression coefficient for the SNP *j*, and the pattern continues for more cohorts.

We are interested in testing the replication null hypothesis, which is a composite null hypothesis. That is, a SNP *j* is considered replicated only when *α_j_* and *β_j_* are both nonzero and share the same effect direction (if there are two SNPs). In other words, the null hypothesis includes the cases where either *α_j_* or *β_j_* is non-zero as well as the case where they point in different directions. More formally, for two cohorts, we have 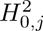 : *α_j_* = 0 ∪ *β_k_* = 0 ∪ {sign(*α_j_*) ≠ sign(*β_j_*)} for SNP *j* [19, 22]. For three cohorts, we have 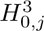 : *α_j_* = 0 ∪ *β_j_*= 0 ∪ *γ_j_*= 0 ∪ {sign(*α_j_*) ≠ sign(*β_j_*) ∪ sign(*α_j_*) ≠ sign(*γ_j_*) ∪ sign(*β_j_*) ≠ sign(*γ_j_*)}.

### Other popular approaches to identify high-quality SNPs

Another popular approach for integrating multiple GWAS to find high-quality SNPs is to perform meta-analysis [1, 9, 13, 14]. However meta-analysis tools such as METAL [50] test the global null hypothesis, which is 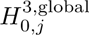 : *α_j_* = *β_j_* = *γ_j_* = 0 for the three-study example. Thus, using meta-analytic type methods does not directly address the scientific question of interest in a replication study. For instance, meta-analysis can identify many SNPs that are highly associated with lung cancer in one study while showing little association in the other two [9]. However, intuitively, such variants would not be considered replicated (1).

The ad-hoc *p*-value threshold approach, e.g. searching for SNPs with *p <* 5 × 10^−8^ in multiple cohorts, can be used to test the replication null. However, this approach is not data-adaptive and does not have good operating characteristics, as we will show. For example, in a relatively small GWAS cohorts where no SNP reaches *p <* 5 × 10^−8^, there will automatically be no replicated findings, even if that cohort has valuable information to contribute (Figure 1).

### Empirical bayes replication testing

We advocate for using a multidimensional empirical Bayes two-group approach [18] paired with a version of the csmGmm [19] for density estimation in replication testing. To the best of our knowledge, this approach has not been used for replication studies in the genetics literature.

In the two-group model, which is popular in many other genomics settings [23], each SNP *j* in each cohort *k* is assumed to possess an unobserved indicator of association status,

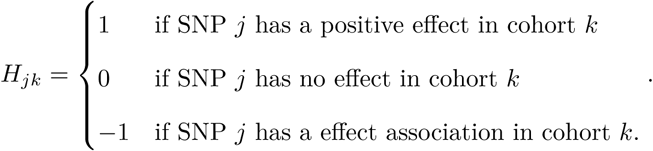

Thus, there is an association group (denoted by 1 and −1) and a no association group (denoted by 0). There are *j* = 1, 2, …, *J* total SNPs and *k* = 1, 2, …, *K* total studies in the analysis.

With two-dimensional studies (*K* = 2), there are *K*^2^ = 9 possible effect configurations [18]. The set of all nine possible effect combinations is given by

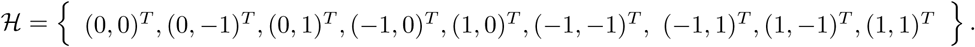

We map each of the nine possible association configurations to a vector **h***^l^*, with *l* = 0, 1*, …L* = 8 in the order given above. Then the two scenarios where the SNP truly possesses a replicated effect are the alternative space, H*_a_* = {**h**^5^ = (−1, −1)*^T^,* **h**^8^ = (1, 1)*^T^* }. The other seven scenarios fall under the null space H_0_. The lfdr [23] for SNP *j* can then be straightforwardly calculated through Bayes’ Theorem as

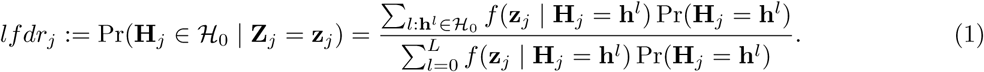

Here, **Z***_j_* = (*Z_j_*_1_*, …, Z_jK_*)*^T^* are the summary statistics for SNP *j* across the *K* studies, and **H***_j_* = (*H_j_*_1_*, …, H_jK_*)*^T^* are the unknown true association configurations for SNP *j* in those studies.

There are many possible densities *f* (·) that can be used to calculate Equation (1) above [18]. We suggest to use a version of the csmGmm model, which addresses the critical issue that empirical Bayes procedures may yield contradictions between Bayesian and frequentist significance rankings [19], for better interpretability. As an example, while other Bayesian methods might declare a SNP with GWAS summary statistics of (*Z_MV P_* = 5.1*, Z_ILCCO_* = 5.2) to be more replicated than a SNP with summary statistics of (*Z_MV P_* = 5.3*, Z_ILCCO_* = 5.4), the csmGmm provably prevents such incongruous conclusions.

### Meta-analysis

We used METAL [50] to perform a meta-analysis on GWAS summary statistics from multiple European ancestry cohorts. A sample size-based method was applied to analyze three different cancers. SNPs with *p <* 1 × 10^−8^ were considered statistically significant. This threshold was selected in accordance with recent lung cancer meta-analysis studies and to maintain a reasonable false discovery rate.

### Simulation studies

We conducted extensive simulation studies to evaluate the power and FDR of various methods in GWAS datasets with the same number of variants as our lung cancer real data analysis (approximately 7 million). Summary statistics were simulated from multivariate normal distributions, and we varied different mean effect sizes and proportions of causal SNPs. We considered both two-cohort (*K* = 2) and three-cohort (*K* = 3) settings, and we performed 100 iterations at each reported setting.

In all simulations, the proportions of causal variants were determined independently. For example, when each dataset contained 1% causal variants, the expected overlap was 0.01% of variants that were causal in both datasets. Also, we used an equal number of positive and negative effects in all cases. A nominal FDR of 0.1 was used for lfdr inference.

### Polygenic risk score construction

Scores were generated with PRSice [31] using clumping and thresholding (LD *r*^2^ = 0.2, 250-kb window, *p*-value threshold = 0.05) and applied to UK Biobank genotypes. Each PRS was standardized to have mean 0 and standard deviation 1. Associations with lung cancer status were assessed via logistic regression adjusted for age, sex, and the first 10 PCs.

### Variant functional annotations

We used genome-wide functional annotations from the FAVOR pipeline [51] to assess the importance of identified variants. Specifically, we considered annotation principal components (aPC) derived from FAVOR for categories including epigenetic function and evolutionarily conserved function. The epigenetics score includes values such as H3K27Ac peaks [52, 53], while the conservation score includes algorithms such as phastCons [54] and PhyloP [55]. The scores are presented on a PHRED scale, so that an aPC epigenetics score of 10 indicates that a SNP ranks in the top 10% of epigenetically active SNPs across the genome. We considered scores of greater than 25 to be high values and those between 10-25 to be medium scores.

## Data Availability

All data produced in the present work are contained in the manuscript.

## Data availability

The R package csmGmm can be used to run the model-based replication analysis described in this manuscript. It is available at https://cran.r-project.org/web/packages/csmGmm/index.html. The code to reproduce of all the tables and figures in this manuscripts are available at https://github.com/yhc0211/Lung-Cancer-Replication-Analysis.

Intermediate files needed to reproduce this work are available at https://odin.mdacc.tmc.edu/rsun3/. For example, the summary statistics from GWAS of lung cancer in the UKB using REGENIE are available here.

## AUTHORS’ CONTRIBUTIONS

YHC: Formal analysis, validation, investigation, visualization, methodology, writing–original draft, writ-ing–review and editing; JB: Writing–review and editing, resources; BRG: Writing–review and editing, resources; RJH: Writing–review and editing, resources; JDM: Writing–review and editing, resources; CIA:Writing– review and editing, resources; SP: Writing–review and editing, resources; AB: Methodology, supervision, investigation, writing–review and editing; RS: Conceptualization, resources, supervision, funding acquisition, investigation, methodology, writing–review and editing

## ACKNOWLEDGEMENTS

This work was supported by National Institutes of Health award R35 GM154843 (R.S.). The funders had no role in study design, data collection and analysis, or preparation of the manuscript.

