## Supplemental Material for "Formal Statistical Replication Analysis in Lung Cancer Genome-Wide Association Studies"

### Supplementary Materials

**Appendix A:** Simulation Results When Using Parameters Similar to ILCCO Real Data

**Appendix B:** Two-Way Replication Analysis Results for ILCCO and UKB

**Appendix C:** Replication Results for Top Sentinel SNPs from ILCCO GWAS

#### Appendix A: Simulation Results When Using Parameters Similar to ILCCO Real Data

| | **number of significant** | | | **Power** | | | **FDR** | | |
|---|---|---|---|---|---|---|---|---|---|
| **% of causal variants** | 0.02 | 0.2 | 1 | 0.02 | 0.2 | 1 | 0.02 | 0.2 | 1 |
| Replication Analysis | 461.20 | 9339.46 | 58676.68 | 0.315 | 0.625 | 0.779 | 0.079 | 0.098 | 0.105 |
| Meta Analysis | 318.19 | 2893.57 | 14299.91 | 0.211 | 0.210 | 0.211 | 0.107 | 0.019 | 0.006 |
| $P < 10^{-8}$ | 0.11 | 1.06 | 5.43 | $8.16 \times 10^{-5}$ | $7.86 \times 10^{-5}$ | $8.05 \times 10^{-5}$ | - | - | - |
| $P < 10^{-6}$ | 2.40 | 25.83 | 126.12 | $1.74 \times 10^{-3}$ | $1.90 \times 10^{-3}$ | $1.87 \times 10^{-3}$ | - | 0.006 | 0.001 |
| $P < 10^{-5}$ | 10.76 | 105.40 | 521.36 | $7.44 \times 10^{-3}$ | $7.77 \times 10^{-3}$ | $7.70 \times 10^{-3}$ | 0.070 | 0.013 | 0.004 |

Supplementary Table 1: Simulation results for two-way replication study under parameters of ILCCO real data. We applied the Expectation-Maximization Algorithm to ILCCO data to estimate the means of summary statistics under the alternative. We also estimated the percentages of causal variants to be approximately 0.2%. We then used the estimated means along with three different causal proportions (0.02%, 0.2%, and 1%) to perform simulations using model-based replication analysis, meta-analysis, and the threshold method. As expected, the model-based replication approach outperforms the other methods in both FDR and power.

### Appendix B: Two-Way Replication Analysis Results for ILCCO and UKB

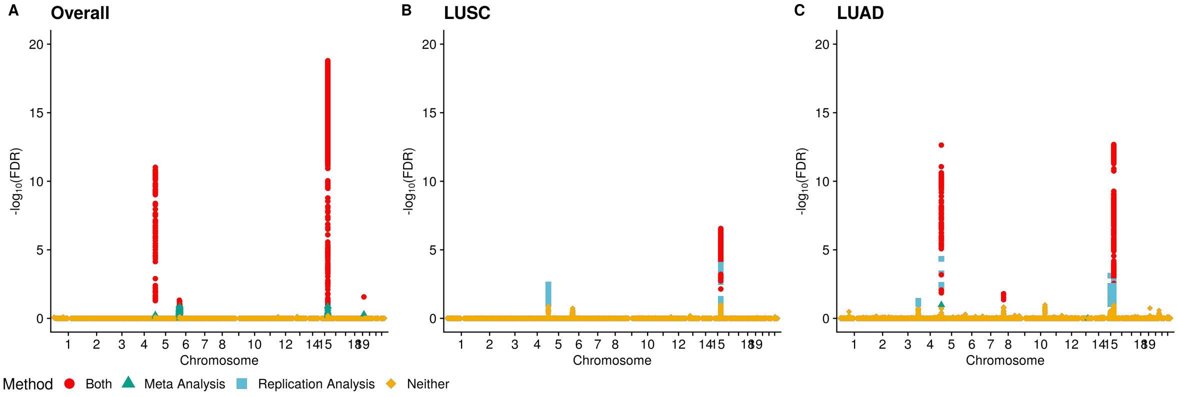


Supplementary Figure 1: *Two-way Replication Analysis Results for ILCCO and UKB.* The y-axis is the $-log_{10}(FDR)$ for SNPs from three different GWAS summary statistics. SNPs prioritized by different methods are highlighted in different colors to illustrate overlap and method-specific signals. Fewer SNPs are replicated in the ILCCO and UKB analysis than the ILCCO and MVP analysis since there is less signal in UKB than in MVP.

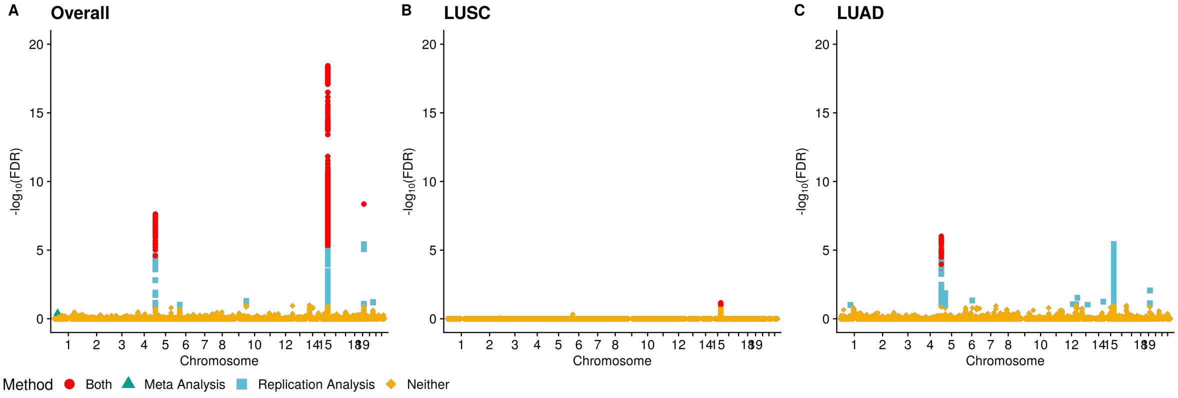


Supplementary Figure 2: *Two-way Replication Analysis Results for UKB and MVP.* The y-axis is the $-log_{10}(FDR)$ for SNPs from three different GWAS summary statistics. SNPs prioritized by different methods are highlighted in different colors to illustrate overlap and method-specific signals. Fewer SNPs are replicated in the MVP and UKB analysis than the ILCCO and MVP analysis since there is less signal in UKB than in ILCCO.

### Appendix C: Replication Results for Top Sentinel SNPs from ILCCO GWAS

| McKay's SNPs (2017) | | | | | Model-based Replication Analysis | | |
|---|---|---|---|---|---|---|---|
| **RS** | **Gene** | **Cancer** | **Chr** | **BP** | **Overall** | **LUSC** | **LUAD** |
| rs71658797 | FUBP1 | Overall | 1 | 77967507 | No | No | No |
| rs6920364 | RNASET2 | Overall | 6 | 167376466 | No | No | No |
| rs11780471 | CHRNA2 | Overall | 8 | 27344719 | No | No | No |
| rs11571833 | BRCA2 | Overall | 13 | 32972626 | No | No | No |
| rs66759488 | SEMA6D | Overall | 15 | 47577451 | No | No | No |
| rs55781567 | CHRNA5 | Overall | 15 | 78857986 | **Yes** | **Yes** | **Yes** |
| rs56113850 | CYP2A6 | Overall | 19 | 41353107 | **Yes** | No | **Yes** |
| rs13080835 | TP63 | LUAD | 3 | 189357199 | No | No | **Yes** |
| rs7705526 | TERT | LUAD | 5 | 1285974 | **Yes** | No | **Yes** |
| rs4236709 | NRG1 | LUAD | 8 | 32410110 | No | No | No |
| rs885518 | CDNK2A | LUAD | 9 | 21830157 | No | No | No |
| rs11591710 | OBFC1 | LUAD | 10 | 105687632 | No | No | **Yes** |
| rs1056562 | AMICA1 | LUAD | 11 | 118125625 | No | No | No |
| rs77468143 | SECISBP2L | LUAD | 15 | 49376624 | No | No | **Yes** |
| rs41309931 | RTEL1 | LUAD | 20 | 62326579 | No | No | No |
| rs116822326 | MHC | LUSC | 6 | 31434111 | No | No | No |
| rs7953330 | RAD52 | LUSC | 12 | 998819 | No | No | No |
| rs17879961 | CHEK2 | LUSC | 22 | 29121087 | No | No | No |

Supplementary Table 2: Top sentinel SNPs identified in ILCCO original GWAS (McKay et. al., 2017) along with replication status when using three-way model-based replication analysis with UK Biobank and MVP data. Some top SNPs have very small p-values in all cohorts. However, many less significant sentinel SNPs do not show evidence of association in all cohorts. In other words, while the most significant SNPs in a GWAS are often replicated, less significant variants are more likely to be false positives, which is an expected result.
