## Extended Data for "Formal Statistical Replication Analysis in Lung Cancer Genome-Wide Association Studies"

### Extended Data Figures and Tables

**Extended Data Figure 1:** Two-way replication analysis results using ILCCO and MVP datasets.
**Extended Data Figure 2:** Comparison of Three-way Model-Based Replication Analysis and Meta-Analysis.
**Extended Data Table 1:** Top SNP from three-way model-based replication analysis for each lung cancer subtype.

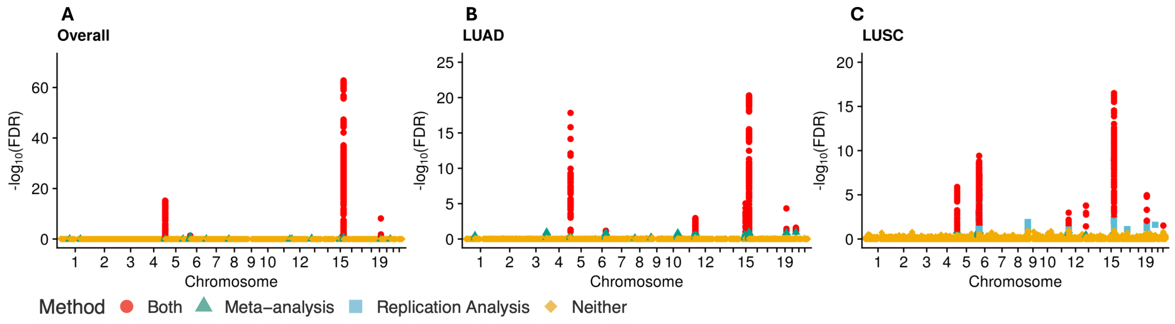


Extended Data Figure 1 *: Two-way replication analysis results using ILCCO and MVP datasets.* The y-axis shows the $-\log_{10}(\text{FDR})$ value from the model-based replication analysis. SNPs are colored red if significant in both model-based replication analysis and meta-analysis, green if significant only in meta-analysis, and blue if significant only in model-based replication analysis. Orange SNPs are not significant in either analysis.

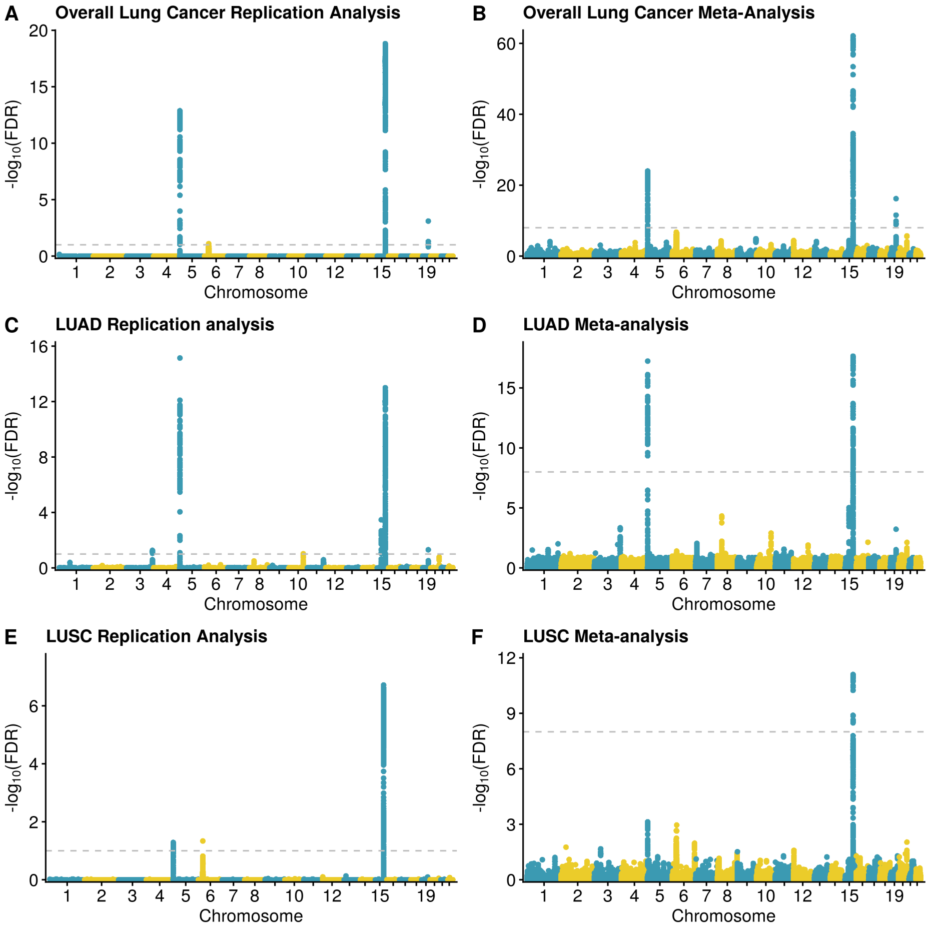


Extended Data Figure 2 : *Comparison of Three-way Model-Based Replication Analysis and Meta-Analysis.* The first column displays results from the replication analysis, and the second column shows results from the meta-analysis. The horizontal dashed gray line indicates an FDR threshold of 0.1, with SNPs above this line considered significant. Odd-numbered chromosomes are colored blue and even-numbered chromosomes gold. Meta-analysis $p$-values were converted to FDR using the Benjamini–Hochberg correction for presentation purposes

| **Chr:Position** | **RSID** | **Gene** | **Cancer** | *P* **ILCCO** | *P* **UKB** | *P* **MVP** | **FDR** |
|---|---|---|---|---|---|---|---|
| 15:78828086 | rs72738786 | HYKK | Overall | $2.25 \times 10^{-99}$ | $1.26 \times 10^{-7}$ | $1.11 \times 10^{-29}$ | $1.52 \times 10^{-19}$ |
| 15:78849918 | rs7173514 | CHRNA5 | LUSC | $6.39 \times 10^{-29}$ | 0.0033 | $3.92 \times 10^{-8}$ | $4.77 \times 10^{-8}$ |
| 5:1287194 | rs2853677 | TERT | LUAD | $6.90 \times 10^{-32}$ | $1.74 \times 10^{-6}$ | $6.08 \times 10^{-9}$ | $7.25 \times 10^{-16}$ |

Extended Data Table 1 : *Top SNP from three-way model-based replication analysis for each lung cancer subtype.* Genomic coordinates (Chr:Position) are based on GRCh37.
